# Knowledge, Attitudes, and Practices regarding Antibiotic Use and Antimicrobial Resistance (AMR) in Nepal

**DOI:** 10.64898/2026.05.27.26354255

**Authors:** Dipesh Thapa, Monika Buda Magar, Thomas Rieger

**Affiliations:** University of Europe for Applied Science

**Keywords:** Antimicrobial resistance, Public health, Knowledge, attitude, practice of AMR

## Abstract

**Background:** Antimicrobial resistance is the world’s silent pandemic. The public knowledge, attitudes, and practices (KAP) about antibiotic usage are strongly related to the growing problem in Nepal.

**Methods:** A cross-sectional descriptive survey was done to 263 respondents. Information on KAP regarding antibiotics, primary healthcare sources, and demography was collected through a questionnaire. To identify health literacy gaps and characteristics that contribute to improper antibiotic use, this study assessed these variables across an age group from 18 to 60 years. Descriptive statistics analysis was performed to analyze the data.

**Results:** The majority of respondents were between the ages of 18 and 39 (85.1%), female (63.1%), and had at least a bachelor’s degree (67.8%). Significant misunderstandings about antibiotics remained, even though 77.6% of respondents correctly recognized antibiotics as effective against bacteria; 44.1% incorrectly believed that antibiotics cure viral diseases, and 87.8% felt that antibiotics should be stopped right away if adverse effects develop. In practice, 52.9% acknowledged quitting antibiotics as soon as symptoms improved, despite 89.4% consulting doctors. Additionally, 43% of respondents said they have taken antibiotics without a prescription, frequently due to pharmacist recommendations (21.67%) and financial or geographical constraints. The main sources of information were doctors (11.07%) and pharmacist-doctor combinations (14.88%), yet 81.8% of respondents said they had never heard of the phrase antimicrobial resistance.

**Conclusion:** There is a significant lack between theoretical understanding and practical application, despite the high levels of fundamental knowledge toward the prohibition of non-prescription sales. Self-medication and early withdrawal are still common inappropriate practices.

It is crucial to implement focused teaching initiatives that highlight the differences between bacterial and viral diseases as well as the risks associated with leftover medicine. It is advised to use digital platforms for younger demographics and to strengthen the role of pharmacists in order to reduce AMR.

## Introduction

Antimicrobial resistance (AMR) is becoming a major worldwide health issue. They are a danger to the effective control of infectious illnesses (Ventola, 2015). The rise and spread of drug-resistant diseases has led to an increase in global rates of illness, mortality, and medical costs (Shankar and Balasubramanium, 2014). One of the main underlying reasons for the rising incidence of AMR is ignorance and misconceptions regarding antibiotics and their misuse (Stanley et al., 2022). Furthermore, antibiotic resistance is also a result of inappropriate antibiotic prescriptions (Nepal et al., 2020). Antimicrobials have been used extensively as growth promoters, treatments, and preventative measures for humans as well as animals for the last 50 years (Llor and Bjerrum, 2014). When some antibiotics or other compounds are used to destroy microorganisms, such as bacteria, viruses, fungi, and parasites that cause infection, enabling the microorganism to survive the drug’s actions, the microorganism may acquire resistance (Sharma et al., 2025). According to estimates, 4.95 million fatalities worldwide in 2019 were attributed to bacterial antimicrobial resistance (AMR), of which 1.27 million were directly related and had no other cause of death (Murray et al. 2022). The knowledge, attitude, and practice (KAP) on antibiotic use and antimicrobial resistance is a comprehensive assessment of people’s behaviors, attitudes, and knowledge on the use of antibiotics and the development of resistance. Behavior includes actual antibiotic-related acts and behaviors, such as self-medication and prescription drug usage (Azim et al., 2023). Numerous variables, such as patient expectations, professional attitudes and practices, misinformation, and ambiguous diagnosis, might lead to the irrational administration of antibiotics (Labi et al., 2018). Multiple reasons, such as the overprescription of antibiotics, empirical therapy, and the selling of medications without a prescription, are contributing to the rise in antibiotic-resistant bacteria. Furthermore, the use of antibiotics for self-medication (SMA) by students, healthcare workers (HCWs), and members of the community has made AMR worse (Mudenda et al., 2022). 60.4% of the 29.19 million people living in Nepal, a low-income Southeast Asian nation, work in agriculture and animal husbandry (Agriculture and Animal Dairy, 2022). According to a recent poll, almost one-third (33.7%) of people in low- and middle-income countries do not completely comprehend antibiotics and how they work (Gualano et al., 2015). Antimicrobial resistance is an issue in poor countries because of insufficient medication limitations, easy access to antimicrobials without a prescription, and a lack of diagnostic tools (Ayukekbong et al., 2017).

## Material and Methods

### Sample design

It is an observational, cross-sectional, prospective, and descriptive study. This survey was done on the Google Forms platform from November 30, 2025, to December 30, 2025.

### Sample size

The sample size of the survey is 263. Data was collected using a questionnaire. The questionnaire comprised 42 questions, and participants were asked to self-complete the survey. All questions were written in English and Nepali, and participation was voluntary and anonymous.

### Inclusion criteria

A random sample method was used to choose the study’s participants. Furthermore, the selected studies contained data on participant demographics, including age, gender, employment experience, and research location. In primary studies that satisfied the following criteria and used standardized questionnaires to conduct surveys of the general public:

Concentrated on general public awareness, attitudes, and practices

Patients, community members, and members of the general public were among the respondents. Both quantitative and mixed-method studies were used; however, the qualitative data given in the mixed-methods research were excluded.

The study was conducted in Nepal.

Described one of the relevant outcomes in the Knowledge, Attitudes, or Practice categories. There were no preset quality requirements that studies had to fulfill to qualify.

### Exclusion criteria

Studies that included participants in aged care or who were older than 60 or younger than 18 were not included, and students enrolled in professional programs or respondents with the power to manage antibiotics were also not included.

### Ethical statement

The French Code of Ethics for Psychologists (2012), the American Psychological Association’s Ethical Principles for Psychologists and Code of Conduct (2017), and the 1964 Declaration of Helsinki and its later modifications (2001) were all applied in this study. Participants received a cover letter outlining the study’s objectives and a guarantee that their information would be kept private.

### Data Analysis

Before analysis, the acquired data were verified for consistency and completeness. Non-response rates were calculated by excluding incomplete questionnaires. Jeffreys’s Amazing Statistics Program (JASP) statistical software was used to analyze the data. Due to their non-normal distribution, the KAP scores on antibiotic use and AMR in this study were demonstrated as numbers, and percentages were used to display the ranked data. The chi-square test was used to investigate any significant relationships between KAP on antibiotic use and AMR, and certain traits and demographics.

## Results

A total of 42 questionnaires were prepared in Google Form, which were disseminated online through various social media platforms, including Facebook and WhatsApp. Among the distributed questionnaires, 263 respondents returned the completed surveys. The majority of the respondents were female (63.1 %) and lived in the urban area (71.5%). The maximum education level is bachelor’s (45.4%), and the age group of 18-28 (45.2%). In addition, a large number of respondents are employed (58.6%), and they are not employed in healthcare (60.8%). It is noted that 12.5% of respondents have at least one chronic disease. Detailed demographic characteristics of the respondents are illustrated in Table 1.

**Table 1:**
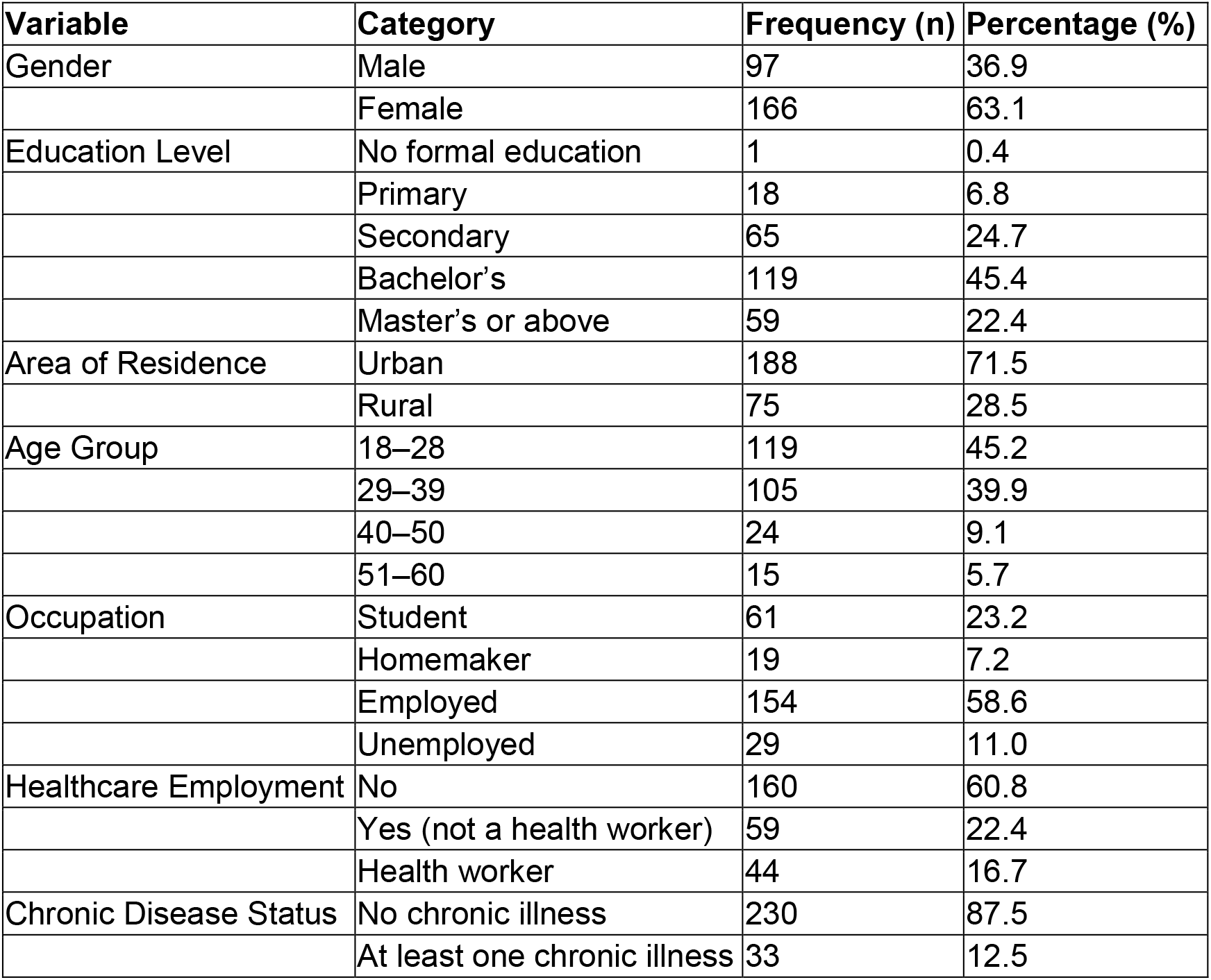
Demographic features of participants.

### Demographic

#### What is your primary source of healthcare?

The survey indicates that public hospitals were the most commonly utilized source of healthcare, accounting for 56.65% of respondents (n = 149). A very small proportion of participants reported relying on traditional healers (1.90%, n = 5). A minority of respondents reported using multiple healthcare sources. The most common combination was public and private hospitals (4.18%, n = 11), followed by public, private hospitals, and pharmacies (2.66%, n = 7) (Table 2).

**Table 2:**
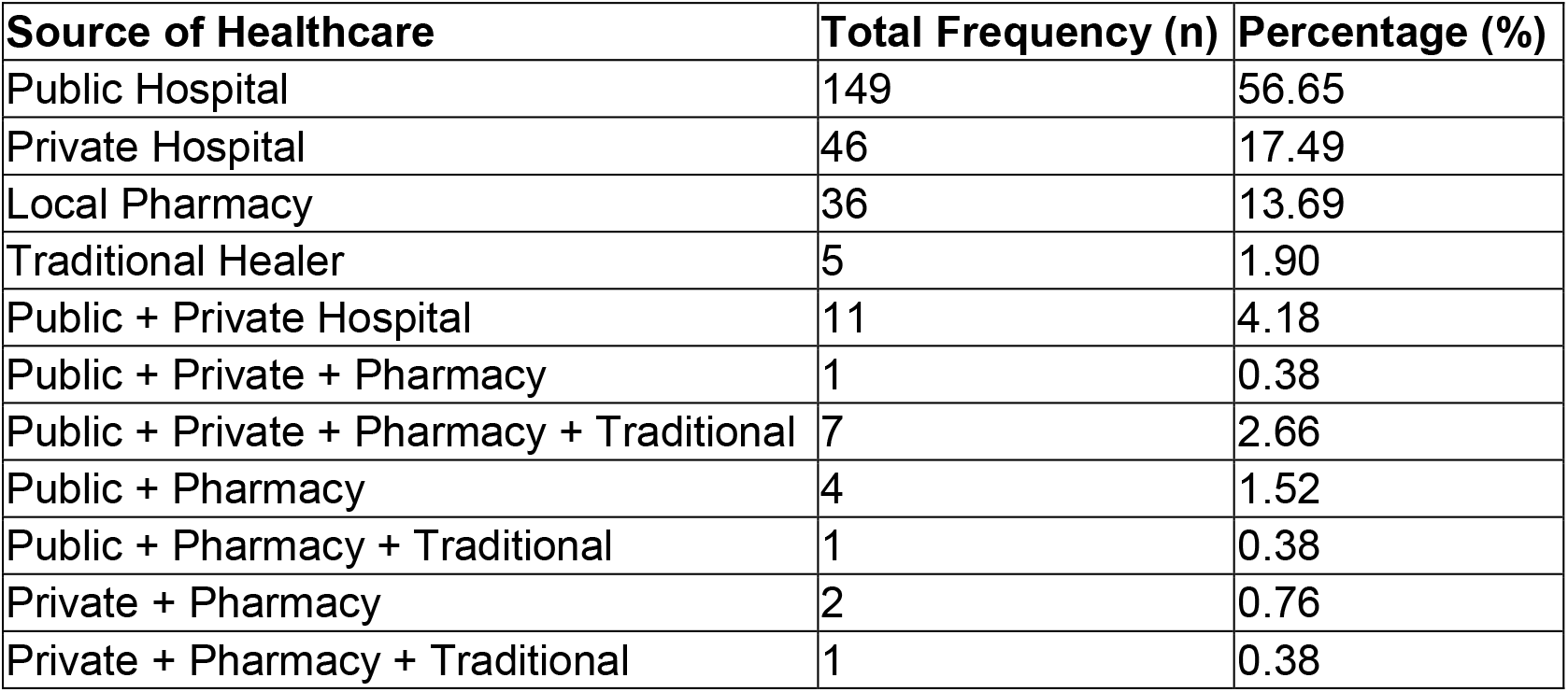
Primary source of healthcare.

### Attitude

The survey on antibiotic use shows strong consensus against self-medication (37.6% and 48.7%). Further study showed that respondents reflect that the high literacy regarding completing an antibiotic course, even if feeling better (33.5% and 55.1%). A high number of participants support prohibiting non-prescription antibiotic sales (27.8 % and 65%), paralleled by this, (27.4 % and 70.3%) of respondents demanded more education and awareness about antibiotic use. While (36.9% and 24 %) of respondents identified antimicrobial resistance as a major problem in my community. It is noted that there is a knowledge gap regarding antibiotics; around 25.1 % of respondents believe antibiotics are necessary for most fevers (Table 3).

**Table 3:** Frequency table of attitude.

| Response | Strongly agree | Agree | Neutral | Disagree | Strongly disagree |
| --- | --- | --- | --- | --- | --- |
| Self-medication with antibiotics is acceptable. | 7 (2.7%) | 10 (3.8%) | 19 (7.2%) | 99 (37.6%) | 128 (48.7%) |
| Antibiotics are necessary for most fevers. | 8 (3%) | 66 (25.1%) | 65 (24.7%) | 92 (35%) | 32 (12.2%) |
| I prefer to get antibiotics directly from the pharmacy rather than seeing a doctor. | 1 (0.4%) | 63 (24%) | 35 (13.3%) | 65 (24.7%) | 99 (37.6%) |
| It is important to finish an antibiotic course even if I feel better. | 145 (55.1 %) | 88 (33.5%) | 17 (6.5%) | 7 (2.7%) | 6 (2.3%) |
| Selling antibiotics without a prescription should be prohibited. | 171 (65%) | 73 (27.8%) | 8 (3%) | 6(2.3%) | 5(1.9%) |
| More education and awareness about antibiotic use are needed. | 185 (70.3%) | 72 (27.4%) | 4 (1.5%) | 2 (0.8%) | 0 |
| Antimicrobial resistance is a major problem in my community. | 63 (24%) | 97 (36.9%) | 86 (32.7%) | 13 (4.9%) | 4 (1.5%) |

### Knowledge

A majority (77.6%) correctly identified that antibiotics are effective against bacteria. Similarly, strong awareness was observed regarding antibiotic resistance, with 89.7% recognizing that misuse can lead to resistance and 92% acknowledging that completing the full course helps prevent it. Most participants (91.6%) understood that taking antibiotics without a doctor’s prescription is unsafe, and 79.5% rejected the idea that using leftover antibiotics is safe. Additionally, 78.7% were aware that overuse in humans and animals contributes to resistance, while 70.3% knew that resistant bacteria can spread from person to person. However, some misconceptions persist. Notably, 44.1% of respondents incorrectly believed that antibiotics can treat viral infections, and 87.8% agreed that antibiotics should be stopped immediately if side effects occur, indicating a misunderstanding regarding proper treatment adherence. Furthermore, only 55.1% correctly identified that a new antibiotic prescription is needed for each illness (Table 4).

**Table 4:** Analysis table of Knowledge regarding the antibiotics.

|  | Don't know | True | False |
| --- | --- | --- | --- |
| Antibiotics are effective against bacteria | 31 (11.8%) | 204 (77.6%) | 28 (10.6%) |
| Antibiotics can cure infections caused by viruses | 18 (6.8%) | 116 (44.1%) | 129(49%) |
| Taking antibiotics without a doctor's prescription is safe | 4 (1.5%) | 18 (6.8%) | 241 (91.6%) |
| Misuse of antibiotics can lead to antibiotic resistance | 22 (8.4%) | 236 (89.7%) | 5 (1.9%) |
| Stopping antibiotics too soon can lead to antibiotic resistance | 46 (17.5%) | 187 (71.1%) | 30 (11.4%) |
| Using leftover antibiotics from a previous illness is safe | 19 (7.2%) | 35 (13.3%) | 209 (79.5%) |
| Resistant bacteria can spread from person to person | 36 (13.7%) | 185 (70.3%) | 42 (16%) |
| Completing the full prescribed course of medicine helps prevent resistance | 17(6.5%) | 242 (92%) | 4 (1.5%) |
| The overuse of antibiotics in humans and animals contributes to antibiotic resistance | 32 (12.2%) | 207 (78.7%) | 24 (9.1%) |
| Antibiotics should be stopped immediately if a side effect occurs | 14 (5.3%) | 231 (87.8%) | 18 (6.8%) |
| A new antibiotic prescription is needed for every new illness | 34 (12.9%) | 145 (55.1%) | 84 (31.9%) |

### Practice

The survey indicates that 89.4% of respondents consult a doctor before taking antibiotics. In addition, out of the total, only 54.7% of respondents do not follow the doctor’s instructions properly. Although 82.1% of respondents generally complete their prescribed courses. Over half of respondents (52.9%) acknowledged stopping taking antibiotics once they felt better, such behaviour shows a primary driver of antimicrobial resistance. Furthermore, knowing the risk of antibiotics, 43% of respondents have taken antibiotics without a prescription, and a smaller percentage (29.3%) have used leftover medication from a previous illness. It is noted that 52.5% of respondents do not know the proper way to dispose of unused or expired antibiotics (Table 5).

**Table 5:** Analysis table of antibiotic practice.

|  | Yes | No |
| --- | --- | --- |
| Do you consult a doctor before taking antibiotics? | 235 (89.4%) | 28 (10.6%) |
| If you consulted a doctor before taking antibiotics, did you follow the exact instructions? | 119 (45.2%) | 144 (54.7%) |
| Have you ever stopped taking antibiotics once you feel better? | 139 (52.9%) | 124 (47.1%) |
| Do you complete the full course of antibiotics when prescribed? | 216(82.1%) | 47 (17.9%) |
| Have you ever used leftover antibiotics from a previous illness? | 77 (29.3%) | 186 (70.7%) |
| Have you ever taken antibiotics without a doctor's prescription? | 113 (43%) | 150 (57%) |
| I know the proper way to dispose of unused or expired antibiotics | 125 (47.5%) | 138 (52.5%) |

### Reasons for taking antibiotics without a prescription or using leftovers

The most frequently reported reasons for using antibiotics without a doctor’s prescription were pharmacist suggestion and other reasons, each 21.7%, respectively. A mild illness was mentioned by 15.97% of respondents, while 12.55% reported thinking it was the same illness. Distance to the doctor’s office contributed to 6.84% of cases. Notably, the combination of pharmacist suggestion, distance, and saving money accounted for 14.83% of cases. Less common combinations, such as pharmacist suggestion together with mild illness and/or perceived similarity of illness, or distance combined with mild illness, each accounted for less than 1% of respondents (Table 6).

**Table 6:** Frequency of reasons for taking antibiotics without a prescription or using leftovers.

| Reason | Total Frequency (n) | Percentage (%) |
| --- | --- | --- |
| The pharmacist suggested it | 57 | 21.67 |
| The doctor's office is far | 18 | 6.84 |
| The illness was mild | 42 | 15.97 |
| Thought it was the same illness | 33 | 12.55 |
| Others | 57 | 21.67 |
| Pharmacist + Doctor's office far | 3 | 1.14 |
| Pharmacist + Distance + Save money | 39 | 14.83 |
| Pharmacist + Distance + Save money + Mild illness + Same illness | 1 | 0.38 |
| Distance + Mild illness | 1 | 0.38 |
| Pharmacist + Distance + Mild illness | 1 | 0.38 |
| Distance + Mild illness + Same illness | 1 | 0.38 |
| Distance + Others | 1 | 0.38 |
| Save money + Mild illness + Same illness | 1 | 0.38 |
| Save money + Same illness | 1 | 0.38 |
| Pharmacist + Mild illness | 2 | 0.76 |
| Mild illness + Same illness | 2 | 0.76 |
| Pharmacist + Mild illness + Same illness | 1 | 0.38 |
| Pharmacist + Same illness | 2 | 0.76 |

### Source of Information

Respondents reported a variety of sources for health-related information. The most frequently cited single source was the doctor (11.1%), followed by school (4.96%), while pharmacists, family/friends, social media, and campaigns each contributed less than 2% individually. Combination sources were more common, with doctor plus pharmacist being the leading combination (14.88%). Other notable combinations included doctor, pharmacist, and family/friends (4.2%), and doctor, pharmacist, social media, and internet websites (2.29%). The most complex combination, encompassing doctor, pharmacist, family/friends, TV/radio, social media, internet websites, school, campaigns, and others, accounted for 5.73% of respondents (Table 7).

**Table 7:** Frequency of source of information.

| **Source / Combination of Sources** | **Total Frequency (n)** | **Percentage (%)** |
| --- | --- | --- |
| Doctor | 29 | 11.07 |
| Pharmacist | 4 | 1.53 |
| Family/Friends | 4 | 1.53 |
| TV/Radio | 1 | 0.38 |
| Social Media | 3 | 1.15 |
| Internet Websites | 2 | 0.76 |
| School | 13 | 4.96 |
| Campaigns | 2 | 0.76 |
| Others | 3 | 1.15 |
| Doctor + Pharmacist | 39 | 14.88 |
| Doctor + Social Media | 1 | 0.38 |
| Doctor + Internet Websites | 4 | 1.53 |
| Doctor + School | 1 | 0.38 |
| Doctor + Others | 1 | 0.38 |
| Family/Friends + Others | 1 | 0.38 |
| Social Media + Internet Websites | 1 | 0.38 |
| Internet Websites + School | 2 | 0.76 |
| Internet Websites + School + Campaigns | 1 | 0.38 |
| Doctor + Pharmacist + Family/Friends | 11 | 4.20 |
| Doctor + Pharmacist + Social Media | 5 | 1.91 |
| Doctor + Pharmacist + Internet Websites | 3 | 1.15 |
| Doctor + Pharmacist + School | 5 | 1.91 |
| Doctor + Pharmacist + Campaigns | 2 | 0.76 |
| Doctor + Family/Friends + Campaigns | 1 | 0.38 |
| Family/Friends + TV/Radio + Social Media | 1 | 0.38 |
| TV/Radio + Social Media + Internet Websites | 1 | 0.38 |
| Doctor + Pharmacist + Family/Friends + Social Media | 3 | 1.14 |
| Doctor + Pharmacist + TV/Radio + Social Media | 2 | 0.76 |
| Doctor + Pharmacist + Internet Websites + School | 5 | 1.91 |
| Doctor + Family/Friends + Social Media + School | 1 | 0.38 |
| Doctor + Pharmacist + Family/Friends + TV/Radio + Social Media | 2 | 0.76 |
| Doctor + Pharmacist + Family/Friends + Social Media + Internet Websites | 4 | 1.53 |
| Doctor + Pharmacist + Social Media + Internet Websites | 6 | 2.29 |

| Source / Combination of Sources | Total Frequency (n) | Percentage (%) |
| --- | --- | --- |
| Doctor + Family/Friends + TV/Radio + Social Media + Internet Websites | 2 | 0.76 |
| Doctor + Pharmacist + Family/Friends + TV/Radio + Social Media + Internet Websites | 2 | 0.76 |
| Doctor + Pharmacist + Social Media + Internet Websites + School | 7 | 2.67 |
| Doctor + Pharmacist + Social Media + Internet Websites + Campaigns | 1 | 0.38 |
| Doctor + Pharmacist + Family/Friends + TV/Radio + Social Media + Internet Websites + School | 7 | 2.67 |
| Doctor + Pharmacist + Social Media + Internet Websites + School + Campaigns | 4 | 1.53 |
| Doctor + Pharmacist + Family/Friends + TV/Radio + Social Media + Internet Websites + School + Campaigns | 17 | 6.49 |
| Doctor + Pharmacist + Family/Friends + TV/Radio + Social Media + Internet Websites + School + Campaigns + Others | 15 | 5.73 |

### Demographic

The study’s demographic distribution offers crucial background information for analyzing the patterns of knowledge, attitudes, and behaviors related to antibiotic usage and antimicrobial resistance (AMR). The survey showed females comprising the majority compared to males. Similar to studies by Wirnitzer et al. (2021), this dominance of female respondents is observed as a common pattern in health-related surveys. Due to improved internet access and digital literacy, younger, urban, and more educated populations tend to be over-represented in digital KAP studies carried out in Nepal, India, and Malaysia (Bawazir et al., 2025). The sample showed a gender-based disproportionate distribution: 166 respondents were female, which is 63.1% of the total respondents. Around 97 respondents were male, making up 36.9% of the sample as a whole. Only 28.5% of respondents in the current study lived in rural areas, while the majority of respondents (63.1%) were female and lived in metropolitan areas (71.5%). The study revealed that most respondents demonstrated a moderate level of knowledge regarding antibiotics. Almost half of the participants agree that improper antibiotic use could contribute to AMR (Sharma, G. et al., 2025). This can be due to the gap in knowledge regarding antibiotics among populations from different disciplines, as other studies, while this study covered a diverse background of populations. The knowledge gap might be due to the different backgrounds, such as education, employment, or household. The survey shows that the gender and demographic background factors can play a vital role in awareness of antibiotic use.

The age distribution indicates that the younger age group of 18-28 and 29-39 showed strong participation. This young age group engagement might be due to the strong engagement with technology (Carter et al., 2016). This statistical data showed that the young age group population is exposed to digital health information. The study showed that the 45.6 % have a bachelor’s degree. This pattern aligns with findings from similar online KAP studies conducted in Malaysia, the UAE, Bangladesh, and India, where the majority of participants had tertiary education (Fahim et al., 2023; Ved et al., 2021). Higher education levels are generally associated with improved health literacy and awareness, which may positively influence knowledge and attitudes toward antibiotic use. The majority of the population lives in the urban area, which might have the benefit of better healthcare services, medical professionals, and health-related information. Previous studies have shown that rural residents often face barriers such as limited healthcare infrastructure, transportation challenges, and fewer healthcare professionals, which restrict access to reliable medical information (Douthit et al., 2015; Anderson et al., 2015). The socioeconomic factors, like education, played a significant role in the knowledge and practice related to antibiotics. The majority of the respondents are employed (58.6%). In addition, this reflects the easy access to health care services like doctors. Studies have shown that disadvantaged groups, particularly those in rural areas or with limited resources, face greater challenges in obtaining appropriate medical care and prescriptions (Edessa et al., 2025). Factors such as inadequate healthcare infrastructure, geographic inaccessibility, and limited availability of healthcare professionals further exacerbate these challenges.

### Knowledge

All the respondents showed a moderate level of knowledge regarding antibiotic use; more than half of the respondents have proper knowledge that antibiotics are effective against bacteria (p > 0.05). This result is consistent with research showing that most populations have a very high level of basic knowledge about antibiotics as antibacterial agents (Gualano et al., 2015). However, some misconceptions regarding antibiotics, which can cure infections caused by viruses like colds and flu, are more 63% which they believe antibiotics can cure. Since more than half of the participants think that antibiotics can treat viral infections (such as the flu and cold), it was found that there is a knowledge gap in the population. While most of the respondents believe that antibiotics without a prescription are unsafe. The female gender shows a high level of awareness regarding the unsafe use of antibiotics (p=0.03). The study conducted by Ajie et al. showed that there is a huge knowledge and practice gap (Ajie et al., 2018; Gladys et al., 2025). Awareness of antimicrobial resistance was moderate as more than half of the respondent which are female reconized that misuse and over use can contribute to the resistance, meanwhile the natural process of self healing, pressure on prescriber, storage of emergency supplies which patients cannot wait, this can create the misuse of antiobiotic (Basu et al., 2018; Shehadeh et al., 2012). Knowledge regarding treatment adherence was also inconsistent, as many participants believed it was acceptable to discontinue antibiotics once symptoms improved, despite evidence that premature discontinuation contributes to resistance (Al-Shibani et al., 2017). Likewise, most of the respondents have knowledge that completing the full course of antibiotics. The leftover antibiotics, which are especially old, are from the age of 40-60 years, indicating a risky have misconception. Furthermore, a considerable proportion of participants incorrectly believed that antibiotics should be stopped immediately if side effects occur, reflecting limited functional health literacy. Knowledge regarding the transmission of resistant bacteria was variable, with younger respondents showing greater uncertainty, while most participants correctly identified the role of antibiotic overuse in humans and animals in contributing to resistance, consistent with global findings (Gualano et al., 2015). Finally, although most respondents understood that a new antibiotic prescription is required for each illness, frequent antibiotic users demonstrated lower levels of understanding, suggesting that repeated exposure may reinforce inappropriate behaviors rather than improve knowledge.

### Attitude

Most of the respondents who accept the strong disagree with the 86.3% self medication (disagree and strongly disagree). This response aligns with global evidence indicating that awareness may be high, but self-medication practice of antibiotics still occurs due to accessibility, cost, and prior experience (Aslam et al., 2020; Simegn and Moges, 2022). Furthermore, 28.1% of respondents responded that antibiotics are necessary for most fevers (strong/strongly agree). This reflects the there is a misconception that participation in the belief that the fevers are similar to bacterial infections (Napolitano et al., 2013; Lim et al., 2021). Most of the respondents show a positive result towards finishing the antibiotic course, which is 88.6% (agree/strongly agree). This shows that there is a high level of awareness regarding the effect of antibiotics left over on health. Similarly, there is mass support for the sales of antibiotics without a prescription being prohibited (92.8%), which shows that the policy for the sales of antibiotics should be reformed or implemented properly. Respondents are also demanding proper awareness campaigns and education about the use of antibiotics (97.7%). However, more than half of respondents, 60.9% (agree/strongly agree), suggest that antibiotics are community problems.

### Practice

There is a huge concern about the arrangement consultation behavior and self-medication. A high number of respondents (89.4%) reported that they consult a doctor before taking antibiotics, which indicates that they have health care-seeking behaviour among the population. Despite this, only 45.2 % of respondents reported that they follow the exact instructions given by doctors, which may reflect a gap in provider-patient communications as noted by the previous studies (Yin et al., 2022). Furthermore, as we noticed that 52.9% of respondents reported stopping antibiotics once they felt better, females are more likely to complete treatment. This premature discontinuation is a critical concern, as it contributes to treatment failure and antimicrobial resistance, and aligns with findings that patients often equate symptom relief with recovery (Yin et al., 2021). Even though 82.1% of respondents showed that they completed a full course of antibiotics. The use of leftover antibiotics was noticed by 29.3% of respondents, which reflects a risky and common behaviour of self-medication practice. Furthermore, 43% of respondents conformed to taking antibiotics without a prescription, which indicates practices are widespread and culturally normalized rather than limited to specific groups (Mahmoudi, 2022). There is less proper practice of proper disposal of unused or expired antibiotics, which is 47.5% of respondents, which indicates females have more awareness than males.

### Source of information

From this survey, the frequently used antibiotics were from pharmacist suggestion and other unspecified factors, each response of 21.67%, indicating that due to a lack of proper medical practice, they primarily rely on the pharmacy. In addition, 15.97% and 12.55% respondents, respectively, believe that they are using antibiotics for mild illness and similar illnesses, which indicates that they are self-diagnosing illnesses due to the influence of the internet. Due to the lack of proper health care facilities within the area, 6.84% of respondents reported that the distance to the doctor’s office is far. A combination of factors like pharmacist advice, distance, and cost saving was noted as a high proportion of 14.83% respondents. These findings are consistent with previous studies demonstrating that convenience, economic constraints, and reliance on informal healthcare providers significantly contribute to self-medication practices (Awad and Aboud, 2015).

The report suggests that multiple sources of information are available in society, and there are gaps in awareness, which form the need for proper coordination and multiple levels of strategies to improve the knowledge of antimicrobial resistance and its uses. The wide range of antibiotic-related information shows that doctors (11.07%) circulate more information than other sources, followed by school education (4.96%). Education institutes like colleges, universities, and schools, as well as awareness campaigns, were mostly commonly noted by young generations, and they may help in shaping health literacy (Bhusal et al., 2021). Despite this diversity, awareness of antimicrobial resistance (AMR) remained low, with 81.8% of participants reporting no prior knowledge, consistent with findings from other studies in Nepal (Acharya and Wilson, 2019). Healthcare professionals, particularly doctors, were identified as the most trusted source of information, aligning with international findings that emphasize their central role in patient education (European Commission, 2022). Although pharmacies are more widely available in every place, they are less noted as a primary source of information, but they can play a significant role in spreading the awareness of antibiotics. Digital platforms, including the internet and social media, were also important sources, particularly among younger and more educated individuals, though their reliability remains variable (Semenova et al., 2024).

## Conclusion

This survey suggests that there is a significant gap between knowledge, attitudes, and practices regarding antibiotic use and antimicrobial resistance among the public. Although there is a misconception of antibiotics as self-medication, viral infections, and treatment. Education plays a crucial role in knowledge, while, in the meantime, premature discontinuity and the use of leftover antibiotics. Information sources, like health care professionals, play a vital role. Thus, this suggests that the proper awareness, education, and stricter regulation are needed to promote rational antibiotic use.

## Data Availability

All data produced are available online at

## Author contributions

Monika was responsible for Introduction, Methodology: Questionnaire Preparation, Results Analysis, Chapter V (Discussion), References, and Conclusion.

Dipesh was responsible for the Abstract, References, Methodology Design and Questionnaire Preparation, Results Analysis and Interpretation, Discussion, and Conclusion.

## Ethical approval

This study involved an oral survey among voluntary participants. Verbal informed consent was obtained from all participants before the survey was conducted. Participant anonymity and confidentiality were maintained, and no personally identifiable information was collected. No animal subjects were involved in this study.

## Reference

1. Acharya, K. P., and Wilson, R. T. (2019). Antimicrobial Resistance in Nepal. Frontiers in medicine, 6, 105.

2. Ajie A.A.D., Andrajati R., Radji M. Factors Affecting the Sale of Non-Prescribed Antibiotics In Jakarta, Indonesia: A Cross-Sectional Study. Int. J. Appl. Pharm. 2018;10:243–247.

3. Al-Shibani N., Hamed A., Labban N., Al-Kattan R., Al-Otaibi H., Alfadda S. Knowledge, attitude and practice of antibiotic use and misuse among adults in Riyadh, Saudi Arabia. Saudi Med. J. 2017;38:1038–1044.

4. Anderson TJ, Saman DM, Lipsky MS, Lutfiyya MN. A cross-sectional study on health differences between rural and non-rural US counties using the County Health Rankings. Bmc Health Services Research. 2015;15.

5. Aslam, A., Gajdács, M., Zin, C. S., Ab Rahman, N. S., Ahmed, S. I., Zafar, M. Z., and Jamshed, S. (2020). Evidence of the Practice of Self-Medication with Antibiotics among the Lay Public in Low- and Middle-Income Countries: A Scoping Review. Antibiotics, 9(9), 597

6. Awad, A. I., and Aboud, E. A. (2015). Knowledge, Attitude and Practice towards Antibiotic Use among the Public in Kuwait. PLOS ONE, 10(2), e0117910.

7. Ayukekbong J. A., Ntemgwa M., Atabe A. N. The threat of antimicrobial resistance in developing countries: causes and control strategies. Antimicrobial Resistance and Infection Control. 2017;6(1):47–48.

8. Azim, M. R., Ifteakhar, K. N., Rahman, M. M., and Sakib, Q. N. (2023). Public knowledge, attitudes, and practices (KAP) regarding antibiotics use and antimicrobial resistance (AMR) in Bangladesh. Heliyon, 9(10).

9. Basu, S.; Garg, S. Antibiotic prescribing behavior among physicians: Ethical challenges in resource-poor settings. J. Med. Ethics Hist. Med. 2018, 11, 5.

10. Bawazir, A., Bohairi, A., Badughaysh, O., Alhussain, A., Abuobaid, M., Abuobaid, M., Al Jabber, A., Mardini, Y., Alothman, A., Alsomih, F., AlMuzaini, A., and BaHamdan, M. (2025). Knowledge, Attitude, and Practice Towards Antibiotic Use and Resistance Among Non-Medical University Students, Riyadh, Saudi Arabia. International Journal of Environmental Research and Public Health, 22(6), 868.

11. Bhusal, S., Paudel, R., Gaihre, M., Paudel, K., Adhikari, T. B., and Pradhan, P. M. S. (2021). Health literacy and associated factors among undergraduates: A university-based cross-sectional study in Nepal. PLOS global public health, 1(11), e0000016.

12. Carter, R. R., Sun, J., and Jump, R. L. (2016). A Survey and Analysis of the American Public’s Perceptions and Knowledge About Antibiotic Resistance. Open Forum Infectious Diseases, 3(3).

13. Douthit N, Kiv S, Dwolatzky T, Biswas S. Exposing some important barriers to health care access in the rural USA. Public Health. 2015;129(6):611–620.

14. Edessa, D., Kumsa, F. A., Dinsa, G., and Oljira, L. (2025). Inequity in access to medications among communities in Eastern Ethiopia: a decomposition analysis. BMC Health Services Research, 25(1), 816.

15. European Commission. (2022). Special eurobarometer 522: antimicrobial resistance.

16. Gladys, O. U., Gabriella, C. N., Lawal, F. O., Bukola, F. O., Amaechi, V. C., and Augustine, O. N. (2025). Antibiotics stewardship: prevalence, nature, and factors associated with dispensing of antibiotics without prescription among community pharmacists in Nigeria. Journal of pharmaceutical policy and practice, 18(1), 2498927.

17. Gualano, M. R., Gili, R., Scaioli, G., Bert, F., and Siliquini, R. (2015). General population’s knowledge and attitudes about antibiotics: a systematic review and meta analysis. Pharmacoepidemiology and drug safety, 24(1), 2–10.

18. Labi, A. K., Obeng-Nkrumah, N., Bjerrum, S., Aryee, N. A. A., Ofori-Adjei, Y. A., Yawson, E., and Newman, M. J. (2018). Physicians’ knowledge attitudes, and perceptions concerning antibiotic resistance: a survey in a Ghanaian tertiary care hospital. BMC health services research, 18(1), 126.

19. Lim, J. M., Duong, M. C., Cook, A. R., Hsu, L. Y., and Tam, C. C. (2021). Public knowledge, attitudes and practices related to antibiotic use and resistance in Singapore: A cross-sectional population survey. 10.1136/bmjopen-2020-048157

20. Llor, C., and Bjerrum, L. (2014). Antimicrobial resistance: risk associated with antibiotic overuse and initiatives to reduce the problem. Therapeutic advances in drug safety, 5(6), 229–241.

21. Mahmoudi, H. (2022). Assessment of knowledge, attitudes, and practice regarding antibiotic self-treatment use among COVID-19 patients. GMS Hygiene and Infection Control, 17, Doc12.

22. Mudenda, S., Mukela, M., Matafwali, S., Banda, M., Mutati, R. K., Muungo, L. T., … and Chabalenge, B. (2022). Knowledge, attitudes, and practices towards antibiotic use and antimicrobial resistance among pharmacy students at the University of Zambia: implications for antimicrobial stewardship programmes. Scholars Academic Journal of Pharmacy, 11(8), 117–124.

23. Murray, C. J., Ikuta, K. S., Sharara, F., Swetschinski, L., Aguilar, G. R., Gray, A., … and Tasak, N. (2022). Global burden of bacterial antimicrobial resistance in 2019: a systematic analysis. The lancet, 399(10325), 629–655.

24. Napolitano F, Izzo MT, Di Giuseppe G, Angelillo IF (2013) Public knowledge, attitudes, and experience regarding the use of antibiotics in Italy. PLoS One 8: e84177.

25. Nepal, A., Hendrie, D., Robinson, S., and Selvey, L. A. (2020). Analysis of patterns of antibiotic prescribing in public health facilities in Nepal. The Journal of Infection in Developing Countries, 14(01), 18–27.

26. Semenova, Y., Kassym, L., Kussainova, A., Aimurziyeva, A., Makalkina, L., Avdeyev, A., Yessmagambetova, A., Smagul, M., Aubakirova, B., Akhmetova, Z., Yergaliyeva, A., and Lim, L. (2024). Knowledge, Attitudes, and Practices towards Antibiotics, Antimicrobial Resistance, and Antibiotic Consumption in the Population of Kazakhstan. Antibiotics, 13(8), 718

27. Shankar, P. R., and Balasubramanium, R. (2014). Antimicrobial resistance: global report on surveillance 2014. Australasian Medical Journal (Online), 7(5), 237.

28. Sharma, G., Paudel, S., Chalise, A., Sapkota, B., and Marasine, N. R. (2025). Knowledge, Attitude, and Practice on Antibiotic Use and Resistance Among Undergraduates, Pokhara Metropolitan, Nepal. BioMed research international, 2025, 9928264.

29. Shehadeh, M., Suaifan, G., Darwish, R. M., Wazaify, M., Zaru, L., and Alja’fari, S. (2012). Knowledge, attitudes and behavior regarding antibiotics use and misuse among adults in the community of Jordan. A pilot study. Saudi pharmaceutical journal : SPJ : the official publication of the Saudi Pharmaceutical Society, 20(2), 125–133.

30. Simegn, W., and Moges, G. (2022). Antibiotics Self-Medication Practice and Associated Factors Among Residents in Dessie City, Northeast Ethiopia: Community Based Cross-Sectional Study. Patient Preference and Adherence, 16, 2159.

31. Stanley, D., Batacan Jr, R., and Bajagai, Y. S. (2022). Rapid growth of antimicrobial resistance: the role of agriculture in the problem and the solutions. Applied Microbiology and Biotechnology, 106(21), 6953–6962.

32. Ventola, C. L. (2015). The antibiotic resistance crisis: part 1: causes and threats. Pharmacy and therapeutics, 40(4), 277.

33. Wirnitzer, K. C., Drenowatz, C., Cocca, A., Tanous, D. R., Motevalli, M., Wirnitzer, G., Schätzer, M., Ruedl, G., and Kirschner, W. (2020). Health Behaviors of Austrian Secondary Level Pupils at a Glance: First Results of the From Science 2 School Study Focusing on Sports Linked to Mixed, Vegetarian, and Vegan Diets. International Journal of Environmental Research

34. Yin, X., Gong, Y., Sun, N. et al. Prevalence of inappropriate use behaviors of antibiotics and related factors among chinese antibiotic users: an online cross-sectional survey. BMC Infect Dis 22, 689 (2022).

35. Yin, X., Mu, K., Yang, H. et al. Prevalence of self-medication with antibiotics and its related factors among Chinese residents: a cross-sectional study. Antimicrob Resist Infect Control 10, 89 (2021).

